# Characterization of Neonatal Abstinence Syndrome in Arizona from 2010-2017

**DOI:** 10.1101/2020.05.12.20099531

**Authors:** Emery R Eaves, Jarrett Barber, Ryann Whealy, Sara A Clancey, Rita Wright, Jill Hager Cocking, Joseph Spadafino, Crystal Marie Hepp

## Abstract

This studv aimed to characterize the population of newborn infants with neonatal abstinence syndrome (NAS) and mothers who were opioid dependent at the time of giving birth, in Arizona. We analyzed approximately 1.2 million electronic medical records from the Arizona Department of Health Services Hospital Discharge Database to identify patterns and disparities across socioeconomic, ethnic/racial, and/or geographic groupings. In addition, we identified comorbid conditions that are differentially associated with NAS in infants or opioid dependence in mothers. Our analysis was designed to assess whether indicators such as race/ethnicity, insurance payer, marital status, and comorbidities are related to the use of opioids while pregnant.

In this paper, we describe a population of mothers who are opioid dependent at the time of giving birth and infants who experience withdrawal due to opioid exposure in utero. While there have been studies of national trends in this population (see Patrick et al.), regional trends and issues are less well understood. Using data from the Arizona Department of Health Services Hospital Discharge Database, we find that women and infants who are non-Hispanic White and economically disadvantaged, tend be part of our populations of interest more frequently than expected. Additionally, we find that women who are opioid dependent at the time of giving birth are unmarried more often than expected, and we suggest that marital status could be a proxy for support. Finally, we report comorbidities, some of which have not been previously reported, associated with infants who have NAS and mothers who are opioid dependent.

## Introduction

Neonatal Abstinence Syndrome (NAS), also called Neonatal Opioid Withdrawal Syndrome (NOWS), is a consequence of abrupt withdrawal from intrauterine opioid exposure after birth [1-3]. Clinical abstinence symptoms are observed in 60-80% of substance exposed neonates, and include neurological, gastrointestinal, and autonomic complications [1]. Neonates with NAS are commonly treated with morphine and methadone to ease withdrawal symptoms, and occasionally with the addition of phenobarbital and clonidine as adjunctive agents [1, 4, 5]. Chronic opioid exposure in utero occurs in three different contexts: (1) active, untreated addiction to opioids (heroin or prescription opioids); (2) opioids used for chronic pain management; and (3) Medication Assisted Treatment (MAT) such as methadone or buprenorphine during pregnancy [6]. Symptoms commonly used to identify NAS cases include tremors, seizures, convulsions, feeding problems, vomiting, diarrhea, respiratory problems, and other neonatal complications [7]. In this paper, we describe the results of analysis of Arizona Hospital Discharge records from 2010 to 2017 to characterize the population of infants born with NAS and their mothers. We found that additional comorbid conditions were more likely to co-occur with NAS than several of the commonly used diagnostic criteria. Our results suggest a need for better characterization of comorbid conditions in NAS infants and their mothers. Improving understanding in these areas has implications for identification and secondary prevention of NAS.

Current NAS prevention strategies include family planning services, prenatal and preconception medical care, changes in the prescribing of opioid drugs, and routine substance use screenings [8, 9]. There are major inconsistencies in substance use screening and NAS treatment [10, 11] and the American Academy of Pediatrics has called for more similarity and standardization of care for infants with NAS [12]. Standardized screening and treatment also has the potential to improve care for infants with NAS and their mothers who are opioid dependent at the time of giving birth [10, 13]. Better and more reliable criteria for screening and early identification of NAS cases is a key gap in efforts to improve and standardize treatment.

Nationwide, NAS cases are more common in rural than in urban areas [14-16]. Between 2004 and 2013, there was a 7-fold increase in NAS in rural areas alone [17]. In Arizona, where 80% of the population live in mental health professional shortage areas [18], increasing availability of illicit drugs and steady rates of prescription opioid pain reliever use impact the population, including pregnant women [19]. NAS incidence in Arizona was previously reported to be 1.3 per 1,000 births in 1999, and increased to 3.9 per 1,000 births in 2013, a threefold increase [20]. The number of opioid-related deaths in Arizona has increased 74% from 2012 to 2016, resulting in more than 2 deaths per day in 2016 [21]. Consistent with findings that socioeconomic status is a factor in NAS cases [22], Hussaini and Saavedra reported that nearly 80% of NAS cases in Arizona were paid for by Medicaid, especially in the border regions of the state [19].

On June 5, 2017, Arizona Governor Doug Ducey declared a Public Health State of Emergency due to the opioid epidemic [23]. An *Enhanced Surveillance Advisory* went into effect as a first step toward understanding the current opioid situation in Arizona and to collect data to develop best practices for interventions. As part of this advisory, any opioid-related event (opioid-related death, naloxone doses administered, NAS cases, etc.) must be reported to Arizona Department of Health Services within 24 hours [23]. As such, we hypothesized that the number of reported NAS cases would correspondingly increase, giving us an opportunity to perform a comprehensive characterization of the population of infants with NAS and mothers who are opioid dependent. This analysis is the first of its kind in Arizona, and moves toward a better understanding of the population of Arizona infants born with NAS and their mothers. The analysis presented below is focused on the 5.5 years prior to and 1.5 years following the implementation of Arizona’s opioid surveillance policy. While there have been studies of national trends in this population [22], regional trends and issues are less well understood [15, 24]. In this pilot project, we use data from the Arizona Department of Health Services Hospital Discharge Database to characterize the population of infants born with NAS and their mothers in Arizona from 2010 to 2017.

## Methods

### Data Request

Northern Arizona University (NAU) has a data use agreement with the Arizona Department of Health Services (ADHS), allowing researchers an expedited path to access records in the ADHS Hospital Discharge Database and other databases. We submitted a data request to the Human Subjects Research Board (HSRB) at ADHS, to access electronic medical records for all infants who were born and all mothers who gave birth in Arizona from 2010-2017. Notably, Indian Health Services hospitals are not required to report inpatient and emergency department visits to ADHS, so birth events at these hospitals are not captured. The request was approved as public health surveillance, and we additionally submitted a request for determination of non-human subjects research to the NAU Institutional Review Board. Based on the ADHS HSRB’s determination of public health surveillance, NAU IRB determined the research to be non-human subjects research. This final dataset, which was transferred between two secure servers at ADHS and NAU, included the electronic medical records from 643,370 mothers and 663,353 infants. All variables and descriptions included in the final dataset used in this project are included in Table S1.

### Identifying the Population of Interest

The purpose of this study is to characterize the population of newborn infants with NAS and mothers who are opioid dependent at the time of giving birth. We used insurance codes to identify subpopulations of interest within the larger mother and infant dataset. The dataset spans 2010-2017, including both the International Classification of Diseases, Ninth Revision, Clinical Modification (ICD-9-CM, referred to as ICD9) and the International Classification of Diseases, Tenth Revision, Clinical Modification (ICD-10-CM, referred to as ICD10), as the change from ICD9 to ICD10 was required by all healthcare facilities in the United States no later than October 1, 2015. To identify newborn infants with NAS, we used ICD9 and ICD10 codes 779.5 and P96.1, respectively. Similarly, to identify mothers who were opioid dependent at the time of giving birth, we used ICD9 codes 304.00-304.03 and 304.70-304.73 and ICD10 code F11, including all subcategories.

### Healthcare Utilization

To better understand hospital resource utilization and to serve as a proxy for severity of morbidity, we compared length of stay and total charges of the subpopulations to the total population of Arizona mothers who have just given birth and newborn infants. The two variables were compared to each other using linear regression to better understand how well one explains the other, and t-tests were used to determine if the populations of interest had means that were significantly different.

### Demographic Disparities

We conducted chi-square tests to determine if selected subpopulations belonged to specific racial and/or ethnic groups or used particular insurance payers significantly more often than expected. Similarly, we used a chi-square test to determine if mothers who were dependent on opioids at the time of giving birth had certain marital statuses more frequently than expected. Expected proportions were determined from the entire mother or infant datasets (Table 2).

### Geographic Disparities

To identify geographic locations where there were more opioid dependent mothers at the time of giving birth than expected based on the total number of mothers who gave birth, we conducted a chi-square analysis. This analysis was completed for all non-tribal primary care areas in Arizona, aggregated from 2010-2017. A primary care area (PCA) is an area in which most residents seek primary health care from the same place. The Arizona Department of Health Services states that the PCA is meant to represent residents’ “primary care seeking patterns” [25]. In addition, PCAs are aggregated to prevent reidentification of a patient in Arizona while allowing for resolution of population health issues at a scale better than that at which the geographically large Arizona counties provide.

### Associated Comorbid Conditions

In addition to demographic information, each inpatient and emergency department electronic medical record includes up to 26 ICD billing codes, including admitting and principal diagnosis codes. In the case of the selected subpopulations, these codes may include information regarding comorbidities of NAS or opioid dependence. To understand comorbidity association with NAS and opioid dependence, we selected comorbidities for their importance in classifying NAS and opioid dependence as measured by their average minimum depth to the maximal sub-tree in classification random forests [26]. For this, we used the function var.select, with options method=‘md’ and conservative=low’, in the R library package randomForestSRC [27, 28]. Our data present us with an imbalanced classification problem [29], wherein positive cases of NAS or opioid dependence represent a very small minority of cases, with the majority of cases being negative. In such situations, overall classification performance—hence comorbidity selection---is dominated by the majority class, whereas our interest leans, instead, toward correct classification of the minority class. We use the method of balanced random forests [29], as implemented in the function imbalanced.rfsrc in the R library package randomForestSRC [27, 28], to grow balanced classification random forests for NAS and opioid dependence before computing the importance of comorbidities.

## Results and Discussion

NAS is increasing rapidly in Arizona. While opioid overdose in Arizona has been a cause for great concern, with suspected overdoses (n=32,900, ~35 per day) and deaths (n=3,935, ~4 per day) at epidemic levels from June 15, 2017 through January 16, 2020 [23], the large number of infants with NAS born during the same period (n=1,295) also warrants attention. To address this issue, we characterized the population of infants with NAS and mothers who were dependent on opioids at the time of giving birth within the context of the entire population of infants born and mothers who gave birth in Arizona from 2010 through 2017.

### Healthcare Utilization

To determine the impact of maternal opioid use during pregnancy on both newborn infant and maternal morbidity as well as on healthcare utilization we compared hospitalization rates, average length of stay, and total charges of the entire populations versus the populations of interest. During the period of time represented in these data, the rate of newborn infants who have NAS has more than doubled from approximately 34 in 2010 to 88 in 2017, per every 10,000 births. (Fig. 1). Similarly, the rate of mothers who are opioid dependent at the time of giving birth has increased from 19 to 85 per every 10,000 mothers who have given birth (Fig. 1). While reporting for infants was substantially higher than reporting for mothers in 2010, hospitalization rates have evened out over time.

**Figure 1.**
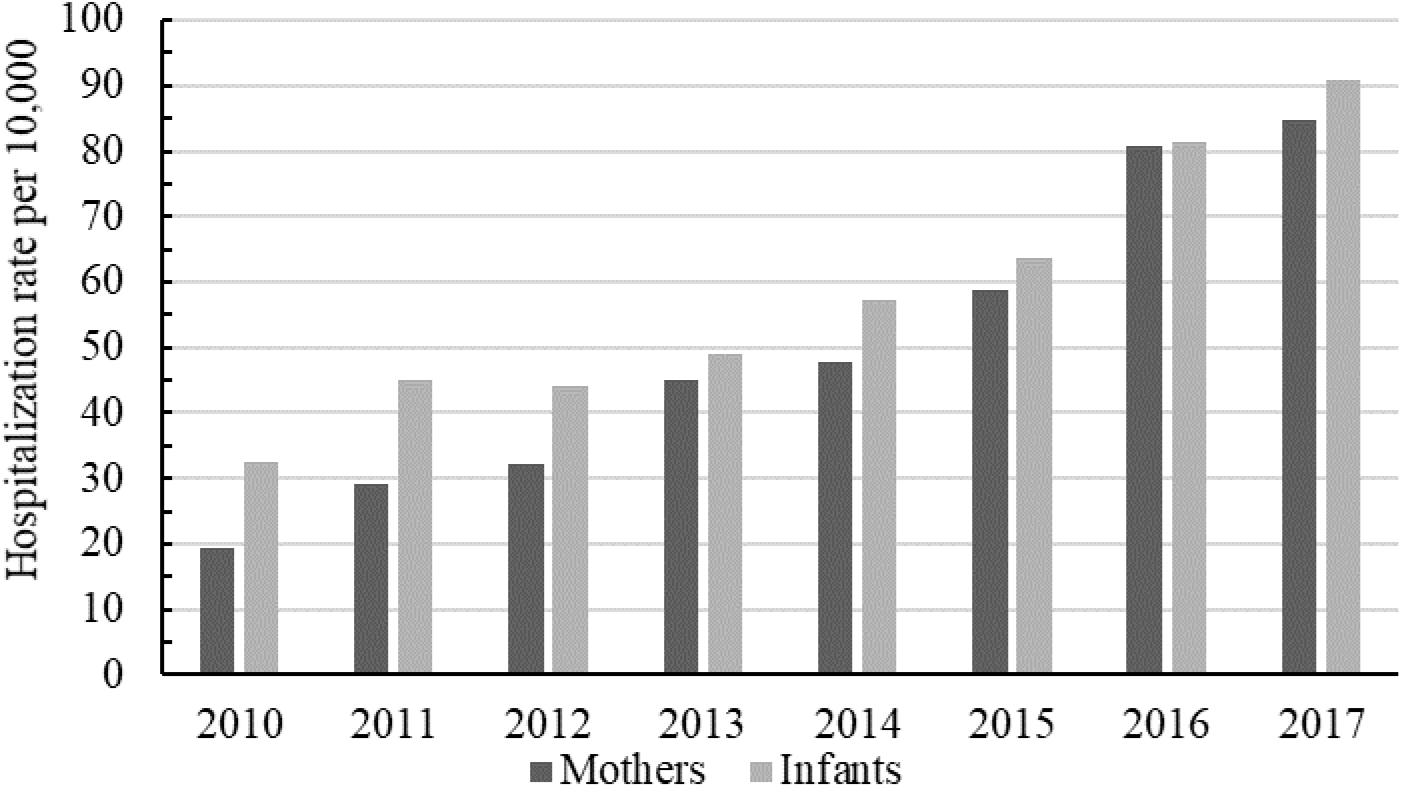
Hospitalization rates, per 10,000 births, in Arizona for mothers who are dependent on opioids at the time of giving birth (dark grey) and newborn infants with NAS (light grey) from 2010 to 2017.

In Arizona, newborn infants with NAS have an average length of stay (mean: 19.71 days, median: 16 days) approximately six times longer than that of all infants (mean: 3.17 days, median: 2 days) (Table 1). Similarly, average total charges are also significantly higher for newborn infants who have NAS (mean: $84,615, median: $49,887) in comparison to all infants (mean: $10,784, median: $3,223). While infants with NAS only compose 0.5% of the total infant population, the total charges associated with infants who have NAS account for 4.5% of all birth related charges ($323,230,298 of $7,153,221,072) from 2010 through 2017. Total charges and length of stay were also significantly higher for mothers who were dependent on opioids at the time of giving birth, however, the differences were modest in comparison to the infant population.

**Table 1.** Comparison of hospital utilization variables in the target versus non-targetpopulations and results of the t-test analyses.

| POPULATION | LENGTH OF STAY (DAYS) | | | TOTAL CHARGES (\$) | | |
| --- | --- | --- | --- | --- | --- | --- |
|  | Mean | Median | p-value | Mean | Median | p-value |
| INFANTS WITHOUT NAS | 3.07 | 2 |  | 10356 | 3209 |  |
| INFANTS WITH NAS | 19.71 | 16 | P<0.0001 | 84615 | 49887 | P<0.0001 |
| NON-OPIOID DEPENDENT MOTHERS | 2.44 | 2 |  | 16801 | 14174 |  |
| OPIOID DEPENDENT MOTHERS | 3.22 | 3 | P<0.0001 | 23674 | 18406 | P<0.0001 |

### Demographic Disparities

Previous studies characterizing infants with NAS and mothers who are dependent on opioids at the time of giving birth in the United States found that these populations were, more often than expected, non-Hispanic white and insured by Medicaid [30, 31]. We additionally examined these demographics to determine if the most heavily impacted populations in Arizona followed national trends. In agreement with previous studies, we found that both infant (Fig. 2) and mother (Fig. 3) populations of interest were, significantly more often than expected, non-Hispanic white, and were Asian or Hispanic/Latino significantly less often than expected. Our target populations were insured by either Medicaid or Medicare significantly more often and by private or military (TRICARE) insurance less often than expected (Figs. 4 and 5).

**Figure 2.**
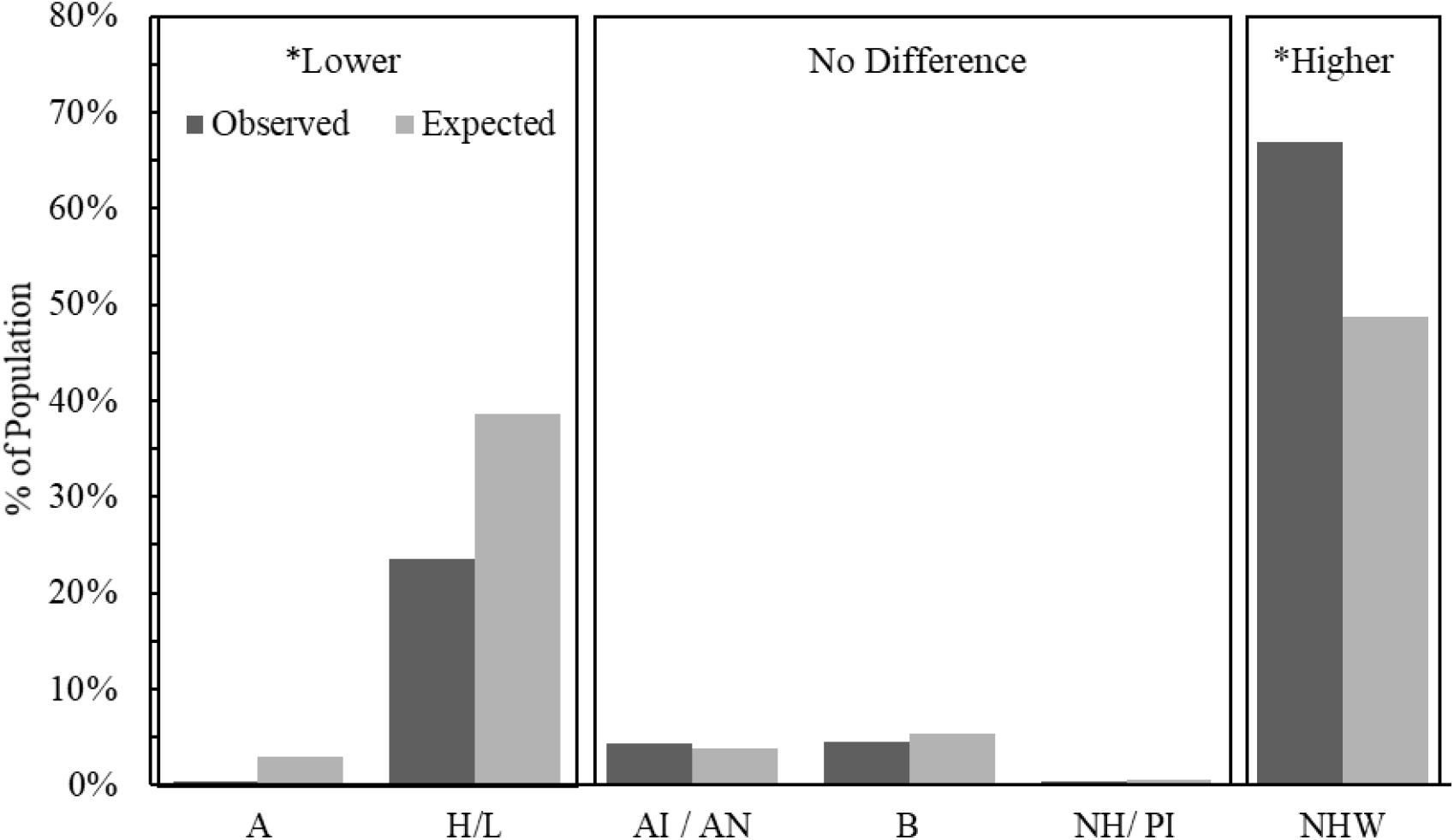
Comparison of the observed versus expected proportions of NAS in each racial/ethnic group of infants. Boxes labelled higher or lower indicate that observed proportions are significantly higher or lower than expected proportions based on a chi-square analysis after post-hoc comparisons that incorporate a Bonferroni correction with six groups (p<0.001/6). A: Asian, H/L: Hispanic or Latino, AI/AN: American Indian or Alaskan Native, B: Black, NH/PI: Native Hawaiian or Pacific Islander, NHW: Non-Hispanic White.

**Figure 3.**
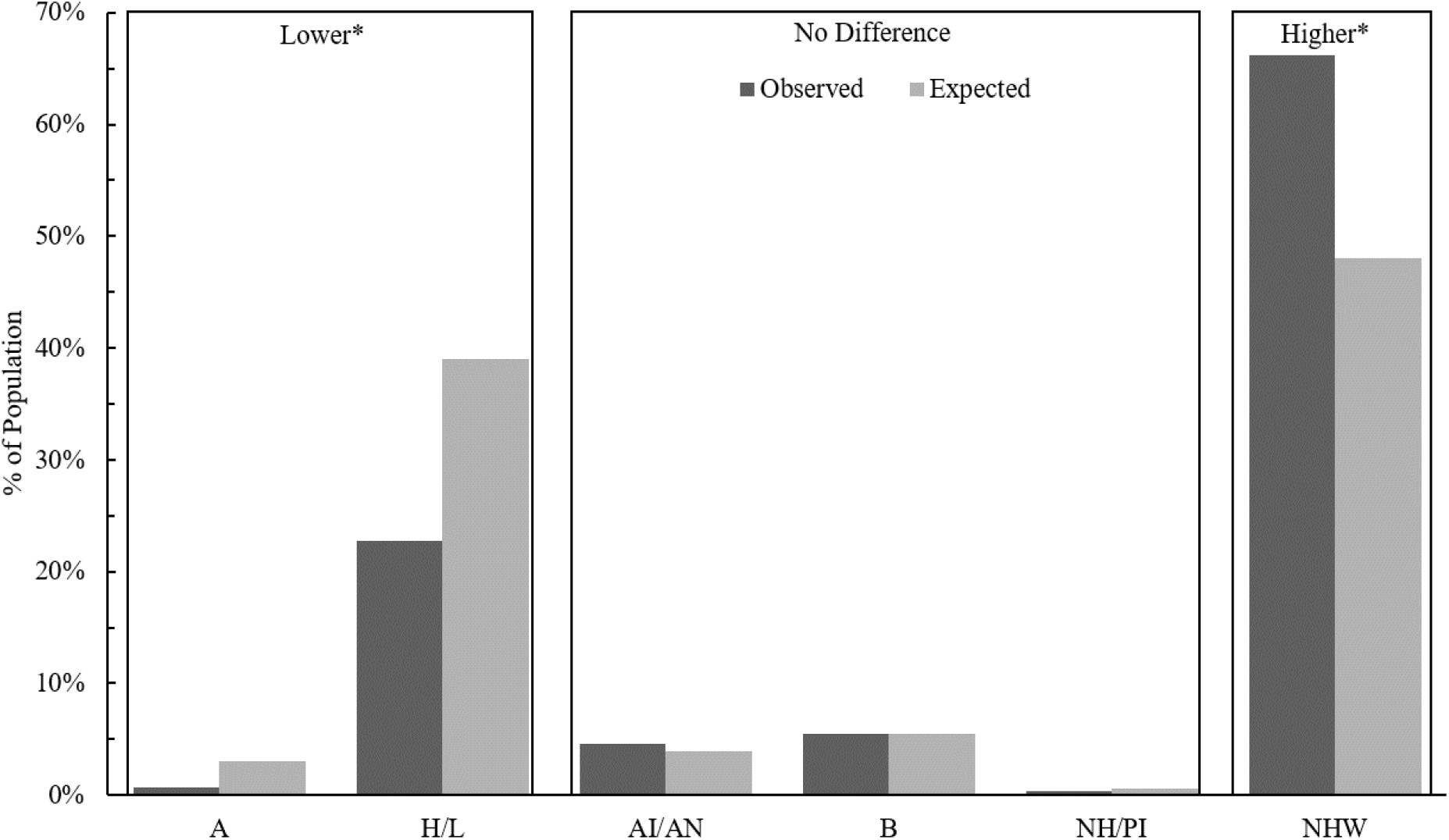
Comparison of the observed versus expected proportions of opioid dependence at the time of giving birth in each racial/ethnic group of mothers. Boxes labelled higher or lower mean that the observed proportions are significantly higher or lower than the expected proportion after post-hoc comparisons that incorporate a Bonferroni correction with six groups (p<0.001/6). A: Asian, H/L: Hispanic or Latino, AI/AN: American Indian or Alaskan Native, B: Black, NH/PI: Native Hawaiian or Pacific Islander, NHW: Non-Hispanic White.

**Figure 4.**
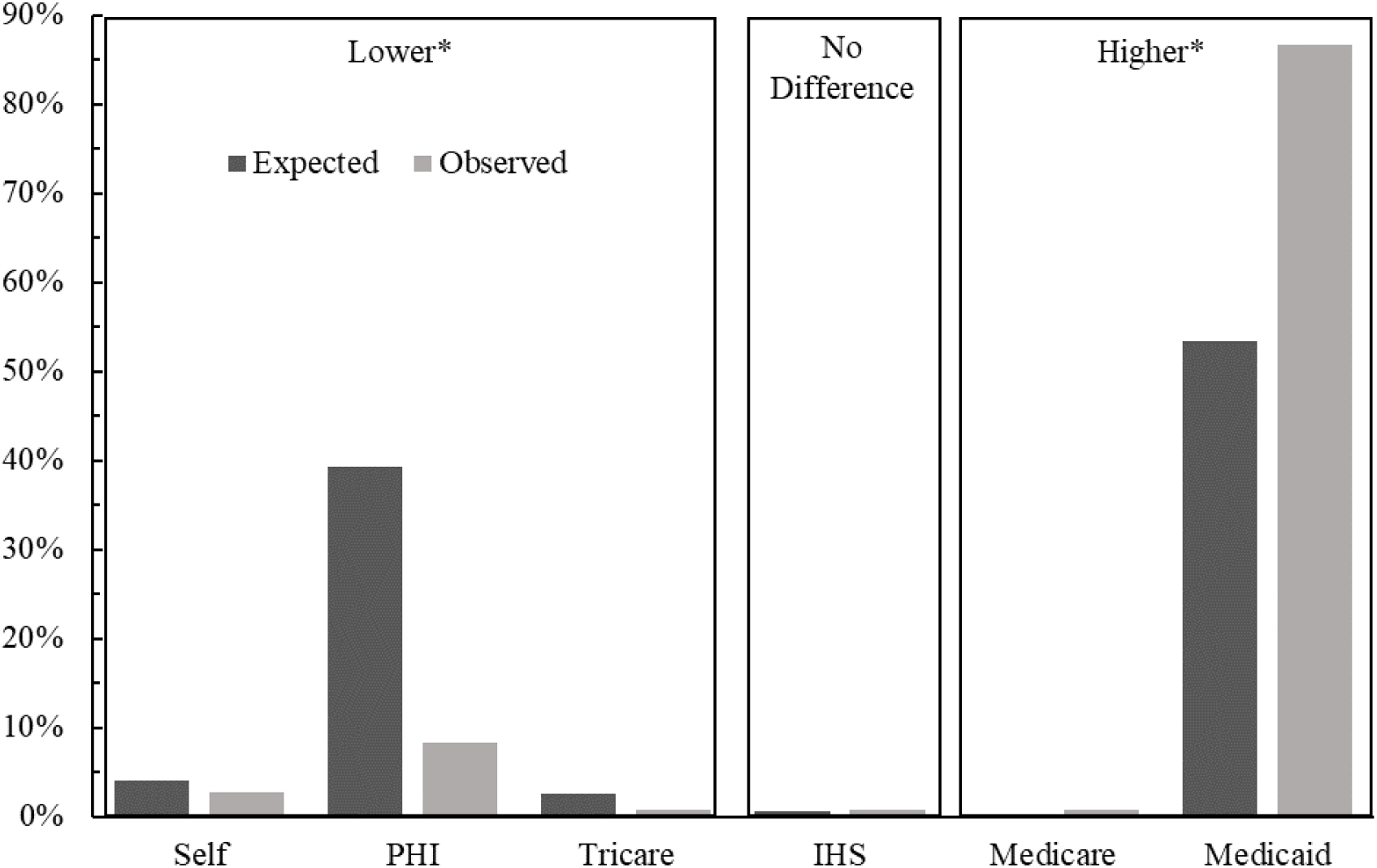
Comparison of the observed versus expected proportions of NAS at the time birth in each payor group utilized for infants. Boxes labelled higher or lower mean that the observed proportions are significantly higher or lower than the expected proportion after post-hoc comparisons that incorporate a Bonferroni correction with six groups (p<0.001/6). PHI: Private Health Insurance, Self: Self Pay, IHS: Indian Health Services.

**Figure 5.**
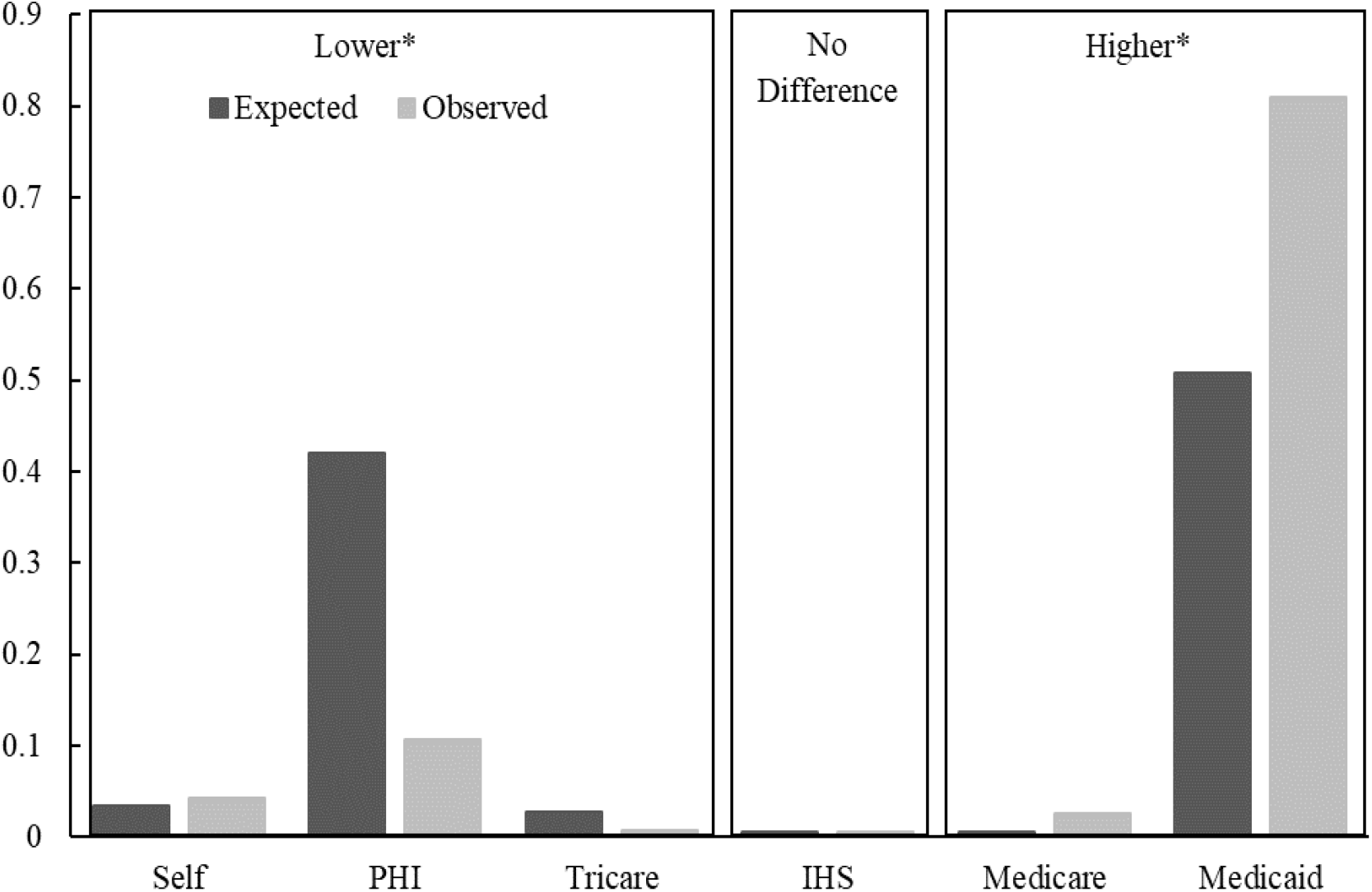
Comparison of the observed versus expected proportions of opioid dependence at the time of giving birth in each payor group utilized for mothers. Boxes labelled higher or lower mean that the observed proportions are significantly higher or lower than the expected proportion after post-hoc comparisons that incorporate a Bonferroni correction with six groups (p<0.001/6). PHI: Private

**Figure 6.**
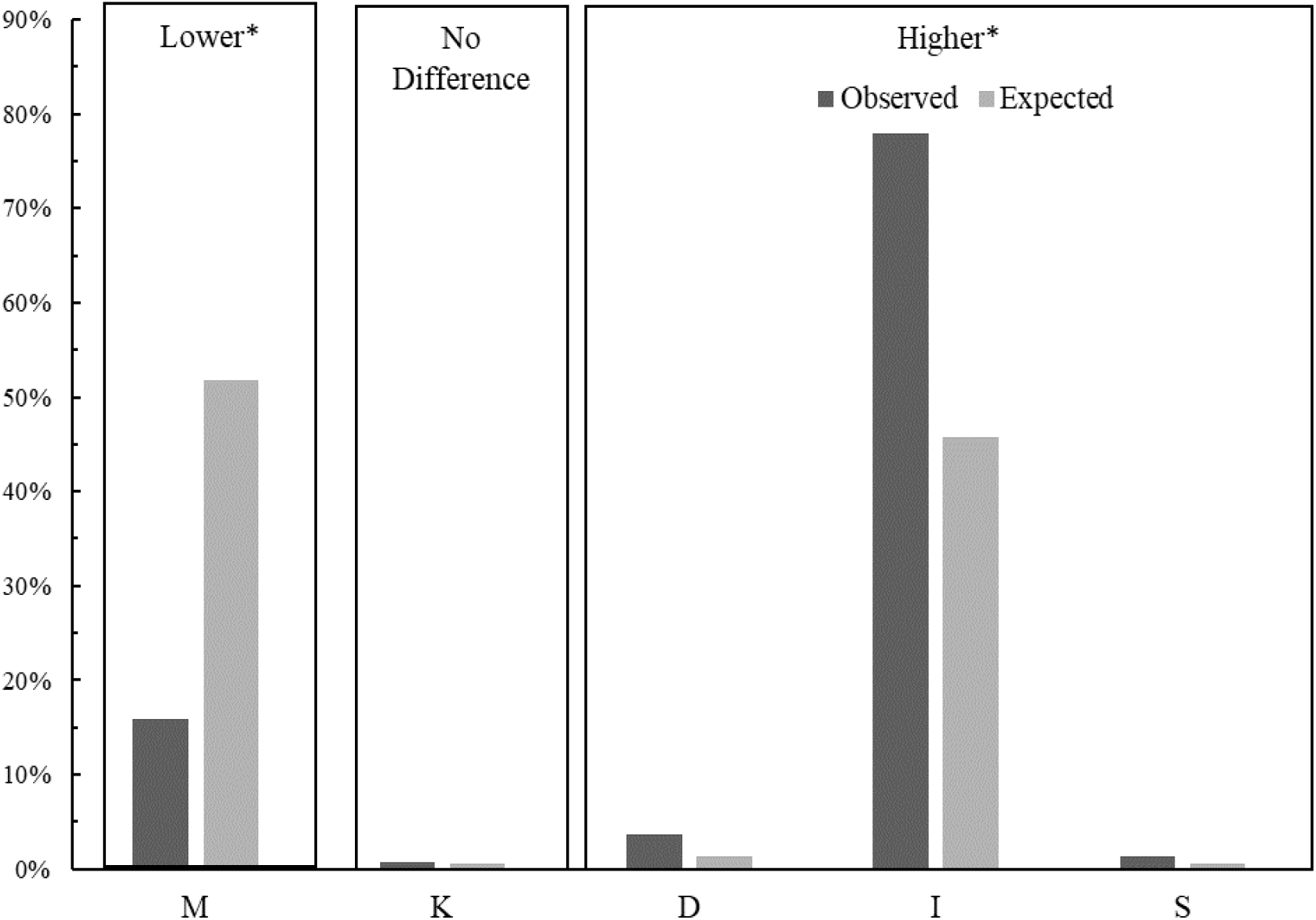
Comparison of the observed versus expected proportions of opioid dependence at the time of giving birth in by maternal marital status. Boxes labelled higher or lower mean that the observed proportions are significantly higher or lower than the expected proportion after post-hoc comparisons that incorporate a Bonferroni correction with six groups (p<0.001/5). M: Married, K: Unknown, D: Divorced, I: Single, S: Separated.

**Figure 7.**
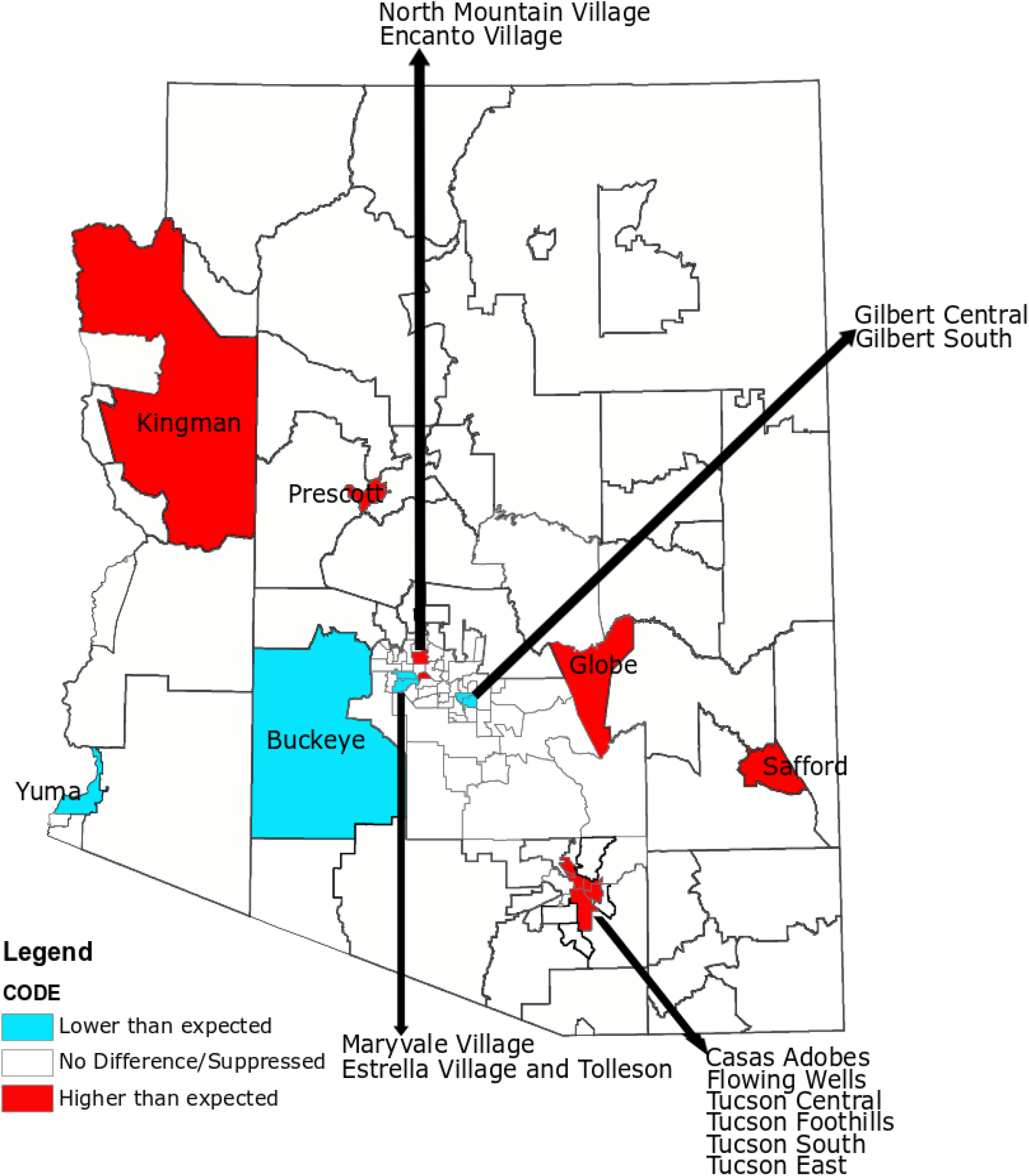
Comparison of the observed versus expected proportions of opioid dependence at the time of giving birth by maternal residential PCA. Red or blue indicates that there is a significantly higher or lower number of mothers using opioids than expected. A map with all PCAs labelled can be found on the ADHS website: https://www.azdhs.gov/documents/prevention/health-systems-development/data-reports-maps/maps/azpca.pdf

We additionally considered maternal marital status and found that women dependent on opioids at the time of giving birth were unmarried significantly more often than expected based on the total population proportions while unmarried women were dependent on opioids significantly less often than expected.

We suppressed data from categories where there were less than 10 mothers (i.e. widowed). Although additional studies are necessary to better understand the association between marital status and opioid use disorder, we suspect that marital status is a proxy for social support. Past studies have found that married individuals are less likely to use illicit drugs [32] and those who participate in substance-abuse treatment programs are more likely to experience positive outcomes [33-36]. Heinz et al. found that that close spousal relationships were a good predictor of reduced cocaine and heroin use in individuals during and after treatment [37]. With the results of these previous studies, our results suggest further investigation into outcomes associated with marital and perhaps other forms of social support when considering opioid use disorder in pregnant women and women of childbearing age.

### Geographic Disparities

Arizona is the 6^th^ largest state in the US by area, but is composed of only 15 counties, where six are among the top 20 geographically largest counties in the United States. The result of a large state being spread into relatively few counties is that distinct human populations are forced into county level estimates which are unlikely to provide a relevant picture of population health. The ADHS has approached this issue by aggregating and reporting population health results for many conditions at the level of PCA. In an effort to identify if and where maternal opioid dependence is clustered, we adopted the ADHS strategy and compared counts across the 126 PCAs that compose Arizona. Within the entire maternal dataset, 25,936 records did not include a PCA, including 75 mothers who were dependent on opioids at the time of giving birth, and these records were not included in the geographic analysis. In addition, we suppressed statistically significant results for PCAs where there were fewer than 10 mothers who were dependent on opioids at the time of giving birth as well as those that are primarily composed of tribal nations. Rather, we have reported those PCAs back to the ADHS for use in their decision processes. An initial chi-square test revealed that opioid dependence among mothers who had given birth significantly deviated from the expected distribution across PCAs. Post-hoc comparisons revealed that there were significantly more mothers who were dependent on opioids residing in the following PCAs than expected: Casas Adobes, Encanto Village, Flowing Wells, Globe, North Mountain Village, Prescott, Safford, Kingman, Tucson Central, Tucson East, Tucson Foothills, and Tucson South. The following PCAs had significantly fewer mothers who were dependent on opioids at the time of giving birth than expected: Buckeye, Estrella Village and Tolleson, Gilbert Central, Gilbert South, Maryvale Village, and Yuma. We are planning future studies to investigate which PCA characteristics may contribute to or mitigate opioid dependence among pregnant women and women of child-bearing age.

### Associated Comorbid Conditions

As mentioned in the Methods section, we used random forests to select comorbidities associated with NAS and opioid dependence. Presence or absence of an ICD9 or 10 code for NAS or opioid dependence was used as the labelled target variable. We analyzed four sets of data for identification of comorbid conditions: Infants with ICD9 codes, Infants with ICD10 codes, Mothers with ICD9 codes, Mothers with ICD10 codes. For infants with NAS (Table 2), we found that, in agreement with previous studies, feeding problems, respiratory distress (transitory tachypnea), and neonatal jaundice commonly co-occurred with NAS [38-40]. We also found that neonatal candidiasis infection and diaper or skin rash (diaper dermatitis) were among top-ranked comorbid conditions. In a recent study of vaginal flora, Farr et al observed significantly higher rates of candidiasis in pregnant mothers receiving medication assisted opioid treatment than in control groups [41]. Diaper dermatitis is a known condition common among infants with NAS, however, it is not considered a reliable diagnostic criterion [42]. Our data suggest that considerably more attention should be paid to potential links between dermatitis and vaginal candidiasis with NAS. To our knowledge there are no studies linking increased rates of neonatal candidiasis with vaginal candidiasis increases among mothers using medication assisted (opioid maintenance) treatment.

**Table 2.** Top ranked ICD9 and ICD10 codes associated with infants who have NAS. ICD10 codes were used for ranking infants admitted after Oct. 1 2015, as well as for any infants born in health care facilities that adopted ICD10 codes prior to Oct. 1, 2015. Conditions are in the ICD9 rank order (see first column), and the corresponding ICD10 and rank of the ICD10 code are listed. The increase from rank 11 to rank 16 was allowed so that all top 10 ICD10 codes could be shown.

| Rank | ICD9 Code | ICD10 Code (and Rank) | Description |
| --- | --- | --- | --- |
| 1 | 779.5 | P961 (1) | Drug withdrawal syndrome in newborn |
| 2 | 779.31 | P92.1-2 and 8-9 (4) | Feeding problems in newborn |
| 3 | 760.72 | P04.49 (2) | Hallucinogenic agents affecting fetus or newborn via placenta or breast milk |
| 4 | 7746 | P599 (5) | Unspecified fetal and neonatal jaundice |
| 5 | 6910 | L22 (3) | Diaper or napkin rash |
| 6 | 770.6 | P22.1 (6) | Transitory tachypnea of newborn |
| 7 | 760.79 | P04.9 (9) | Other noxious influences affecting fetus or newborn via placenta or breast milk |
| 8 | 771.7 | P37.5 (7) | Neonatal Candida infection |
| 9 | V290 | P002 (13) | Observation for suspected infectious condition |
| 10 | V05.3 | Z23 (19) | Need for prophylactic vaccination and inoculation against viral hepatitis |
| 11 | V30.00 | Z38.00 (8) | Single liveborn, born in hospital, delivered without mention of cesarean section (i.e. delivered vaginally) |
| 16 | 745.4 and 745.6 | Q21.1 (10) | Atrial septal defect |

**Table 3.** Top ranked ICD9 and ICD10 codes associated with mothers who are opioid dependent at the time of giving birth. ICD10 codes were used for ranking mothers admitted after Oct. 1 2015, as well as for any infants born in health care facilities that adopted ICD10 codes prior to Oct. 1, 2015. Conditions are in the ICD9 rank order (see first column), and the corresponding ICD10 and rank of the ICD10 code are listed. The increase from rank 12 to rank 62 was allowed so that the top 10 ICD10 codes could be shown.

| Rank | ICD9 Code | ICD10 Code<br>(and Rank) | Description |
| --- | --- | --- | --- |
| 1 | 304 | F11.20,<br>F11.90,<br>F1110(1,3,4) | Opioid type dependence, unspecified |
| 2 | 648.31 | O99.324(2) | Drug dependence of mother, delivered, with or without mention of antepartum condition |
| 3 | 648.41 | O99.344(8) | Mental disorders of mother, delivered, with or without mention of antepartum condition |
| 4 | 649.01 | O99.334 and<br>F17.210(5,6) | Tobacco use disorder complicating pregnancy, childbirth, or the puerperium, delivered, with or without mention of antepartum condition |
| 5 | 338.29 | G89.29(9) | Other chronic pain |
| 6 | V23.7 | O09.30(7) | Supervision of high-risk pregnancy with insufficient prenatal care |
| 7 | 644.21 | O60.14X0<br>(57) | Early onset of delivery, delivered, with or without mention of antepartum condition |
| 8 | 648.91 | No specific<br>conversion | Other current conditions classifiable elsewhere of mother, delivered, with or without mention of antepartum condition |
| 9 | 647.61 | O98.42,<br>O98.511-513,<br>O98.52(31) | Other viral diseases in the mother, delivered, with or without mention of antepartum condition |
| 10 | 300 | F41.9(15) | Anxiety state, unspecified |
| 11 | 664.11 | O70.1(14) | Second-degree perineal laceration, delivered, with or without mention of antepartum condition |
| 12 | 656.51 | O36.5110-130,<br>O36.5910-30<br>(26) | Poor fetal growth, affecting management of mother, delivered, with or without mention of antepartum condition |
| 62 | 305.70-72 | F15.10(10) | Amphetamine or related acting sympathomimetic abuse (unspecified, continuous, or episodic) |

Like the infant analysis, analysis of mothers’ discharge records identified several comorbid conditions that previous studies have found to be associated with opioid dependence during pregnancy. For example, polysubstance use, notably tobacco, alcohol, and stimulants, is more common among pregnant women who use opioids [43]. Tobacco use is particularly prevalent among this population, with estimates of tobacco use as high as 85-90% among pregnant women treated with buprenorphine or methadone [44-46]. Pregnant women using opioids have been found to be more likely to be diagnosed with depression, anxiety, post-traumatic stress disorder, and panic disorder [47], which is also in agreement with our study, where maternal mental disorders are highly ranked. Interestingly, while we found studies that identified increased opioid prescriptions written to women with perineal lacerations [48], to our knowledge no studies have reported an association between perineal lacerations and opioid dependence at the time of giving birth. Epidural anesthesia has been associated with increased perineal laceration [49] and mothers who are opioid dependent at the time of giving birth are not candidates for pain management with opioids [50, 51]. Future research should consider whether the risk of perineal laceration among opioid dependent women is elevated due to pain management practices or other factors.

## DISCUSSION/CONCLUSIONS

Infants with NAS in Arizona and their mothers from 2010-2017 tended to be socioeconomically disadvantaged, non-Hispanic White, and geographically clustered throughout Arizona. In addition, mothers in this group are unpartnered more frequently than expected, which may indicate a relative lack of social support. Unsurprisingly, characteristics of mothers with opioid dependence at the time of birth were closely related to those of infants with NAS. There does not appear to be an increase in reported of maternal opioid dependence from 2016 to 2017. Mothers who were opioid dependent accumulated nearly $8000 more in total charges and stayed a half day longer than those who were not opioid dependent. There does not appear to be a significant age difference between mothers with opioid dependence at the time of birth and those who are not opioid dependent.

Hospital charges for infants born with NAS are paid for with Medicaid or Medicare more often than expected, and less often than expected with private health insurance, military coverage, or self-pay (p < 0.05). On average, infants born with NAS in Arizona stayed in the hospital 6 times longer and have 8 times higher hospital charges than those without NAS. Despite new state mandates to reduce opioid deaths through Enhanced Surveillance on June 5 ^th^, 2017, there does not appear to be a substantial increase in reporting from 2016 to 2017.

Areas with higher than expected rates of NAS in relation to population estimates warrant additional research. In our meetings with stakeholders and investigation of local understandings of areas with higher rates of drug use in the state, observations were consistent with our findings. For example, the Prescott area is known for a proliferation of “sober living houses” in recent years, with so many recovering substance users coming to the area from outside that the trend has been reported in state and national news outlets [52, 53]. Low numbers of medication assisted treatment providers in areas with the highest rates of opioid use and NAS cases are also potentially responsible for higher than expected rates in some, particularly rural, areas of the state [54].

The comorbidity analysis, using supervised machine learning, revealed that diaper dermatitis and jaundice were more importantly associated with NAS than traditional conditions of respiratory distress and irritability. Additionally, to our knowledge, neonatal candidiasis has not been previously associated with NAS. However, our results, coupled with the finding that pregnant women receiving medication assisted opioid treatment are more frequently colonized with *Candida* [41], suggests that it may be important to screen pregnant women receiving medication assisted opioid treatment for candidiasis to develop maternal and neonatal treatment strategies. Our analysis of the Arizona Hospital Discharge Database from 2010 to 2017 suggests a need for better characterization of comorbid conditions in NAS infants and their mothers. Calls to improve standardization in the treatment of NAS infants, and efforts to improve primary and secondary prevention of NAS may benefit from improved understanding of comorbid and frequently co-occurring conditions. Future research should also consider potential interactions between opioid exposure and comorbid conditions in utero.

## Data Availability

Raw data were generated at Arizona Department of Health Services using hospital discharge records. Derived data supporting the findings of this study are available from the corresponding author CMH on reasonable request.

## ACKNOWLEDGEMENTS

This work was supported by a Pilot grant project to Dr. Crystal Hepp as part of the NIH/NIMHD RCMI U54MD012388 to Julie Baldwin and Diane Stearns. The authors acknowledge the contributions of Arizona Department of Health Services staff members Kyle Gardner and Timothy Flood for assistance defining the data request, the Northern Arizona University Information Technology Services for managing data security issues, and the Southwestern Health Equity Research Collaborative Research Infrastructure Core for analytical assistance.

---

**Table S1.** Summary of non-geographic data used in this study by figure. The Mothers^O^ column contains data for all mothers who were dependent on opioids at the time of giving birth. The Infants^NAS^ column contains data for all infants who had NAS. Totals at the end of each category are not the same for all categories due to missing data or because some patients reported atypical categories (e.g. payer was workers compensation or individual was a foreign national).

|  |  | Mothers<sup>O</sup> | Mothers |  | Infants<sup>NAS</sup> | Infants |
| --- | --- | --- | --- | --- | --- | --- |
| Year (Fig. 1) | 2010 | 159 | 82143 |  | 288 | 84772 |
|  | 2011 | 235 | 80350 |  | 370 | 82598 |
|  | 2012 | 263 | 81647 |  | 367 | 83594 |
|  | 2013 | 366 | 81089 |  | 416 | 83032 |
|  | 2014 | 394 | 82382 |  | 486 | 84283 |
|  | 2015 | 474 | 80785 |  | 542 | 82550 |
|  | 2016 | 649 | 80404 |  | 666 | 81827 |
|  | 2017 | 633 | 74570 |  | 675 | 76136 |
|  | Total | 3173 | 643370 |  | 3810 | 658792 |
| Race/Ethnicity<br>(Figs. 2 & 3) | Asian | 21 | 19180 |  | 13 | 19076 |
|  | Hispanic/Latino | 717 | 247391 |  | 889 | 253466 |
|  | American Indian or Alaska Native | 144 | 24556 |  | 165 | 24241 |
|  | Black | 172 | 34460 |  | 170 | 34890 |
|  | Native Hawaiian/Pacific Islander | 11 | 3634 |  | 12 | 3621 |
|  | Non-Hispanic White | 2080 | 304745 |  | 2509 | 316390 |
|  | Total | 3145 | 633966 |  | 3758 | 651684 |
| Insurance<br>(Figs. 4 & 5) | Self | 135 | 21350 |  | 105 | 26093 |
|  | Private | 335 | 264803 |  | 315 | 254420 |
|  | Tricare | 23 | 17032 |  | 30 | 17129 |
|  | IHS | 19 | 3645 |  | 27 | 3713 |
|  | Medicare | 81 | 3335 |  | 27 | 1261 |
|  | Medicaid | 2528 | 318979 |  | 3244 | 345333 |
|  | Total | 3121 | 629144 |  | 3748 | 647949 |
| Marital Status<br>(Fig. 6) | Married | 506 | 333116 |  |  |  |
|  | Unknown | 25 | 4075 |  |  |  |
|  | Divorced | 115 | 8665 |  |  |  |
|  | Single | 2474 | 293773 |  |  |  |
|  | Separated | 44 | 3207 |  |  |  |
|  | Total | 3164 | 642836 |  |  |  |

## Notes

### Competing Interest Statement

The authors have declared no competing interest.

